# Too rare to be random: genetic finding suggests previously unrecognized path of mutagenesis

**DOI:** 10.64898/2026.03.03.26346966

**Authors:** Jonas Böhnlein, Johann G. Maass, Julia Dennig, Sebastian Burkart, Lilian T. Kaufmann, Michelle Brehm, Kirsten Göbel, Annette Kopp-Schneider, Tim Holland-Letz, Katrin Hinderhofer, Maja Hempel, Christian P. Schaaf

**Author notes:** Corresponding author: Christian P. Schaaf. Co-first (Jonas Böhnlein, Johann G. Maass, Julia Dennig).

## Abstract

We report a previously undescribed genotypic configuration identified in twins with *HNRNPU*-related neurodevelopmental disorder. Both twins have two closely spaced mosaic variants on the same allele that never co-occur on any single DNA molecule, resulting in three distinct cell lineages within each individual. We define this genotypic configuration as clustered monoallelic mosaicism (cMoMa). Recognizing the extreme improbability of such a configuration, we systematically explore two potential mechanisms for its origin. We propose that this genotype arises from an asymmetric repair of a single mutational event in an early embryonic cell, yielding divergent outcomes on sister chromatids. Screening of datasets (COSMIC and MosaicBase) uncovered additional cMoMa-like cases, suggesting that the mechanism is not unique to our case, but rather represents a broader, previously unrecognized path of mutagenesis that extends our current definition of mosaicism.

## Introduction

In modern human genetics, most classes of genetic variation are associated with, and can be explained by, established mutagenic pathways. These pathways allow us to understand even highly complex genomic architectures observed in human genomes, including large-scale rearrangements and translocations. However, as genome-wide testing becomes increasingly routine, variants are being identified that cannot be readily explained by the current catalogue of mutagenic mechanisms, suggesting that additional pathways remain to be identified.^1^

## Results

We identified an improbable and previously uncategorized genotype in monochorionic diamniotic (MCDA) twins presenting phenotypes consistent with *HNRNPU*-related neurodevelopmental disorder (OMIM # 617391).^2,3^ Whole-genome sequencing revealed a mutational configuration with two closely spaced, *de novo* 1-bp deletions in *HNRNPU*, each present in mosaic state. Notably, the deletions were mutually exclusive at the molecular level: some reads harbored c.1463del, others c.1466del, but none carried both (Fig. 1 and Supplemental Fig. 1A). Furthermore, long-read sequencing determined that both deletions, while mutually exclusive, are located on the paternal allele (Fig. 1 and Supplemental Fig. 1B). Variant-allele fractions (VAFs) of both pathogenic variants ranged from ∼20-39% in whole-genome analysis from leukocyte-derived DNA. Sanger sequencing of DNA from leukocytes and buccal swabs confirmed both variants and their mosaic distribution (Supplemental Fig. 2). Taken together, each of the twins has three cell types with a wild-type sequence on the maternal allele and either:

**Figure 1:**
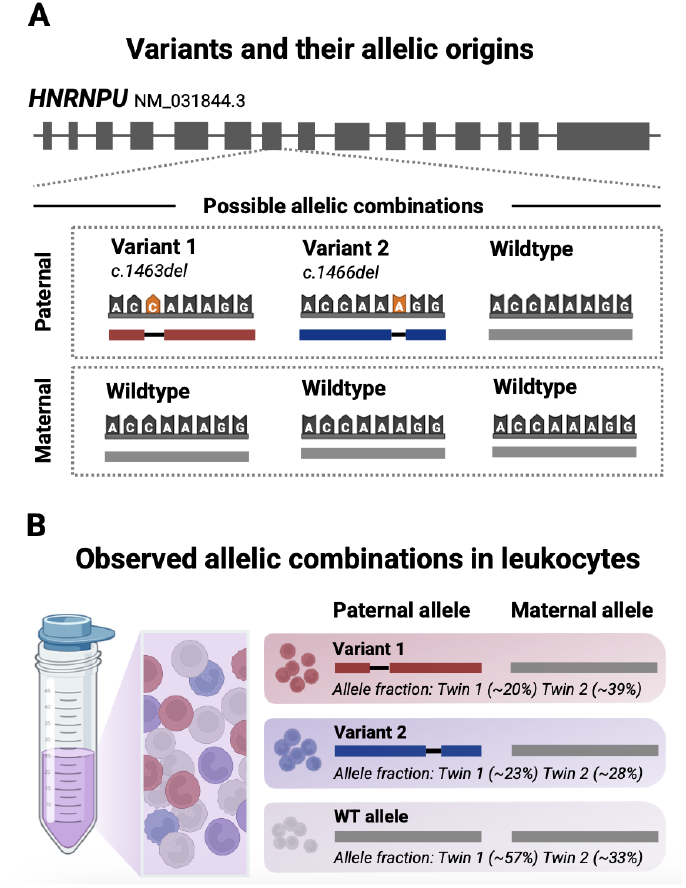
Overview of the genotype discovered in the twins. (A) Schematic of *HNRNPU* pre-mRNA showing three allelic haplotypes. Two closely spaced 1-bp deletions (c.1463del and c.1466del) occur on the paternal allele, with ambiguous positioning of c.1466del. Long-read phasing confirmed both variants on the paternal haplotype and mutual exclusivity at the molecular level. (B) Illustration of the three different allelic combinations identified in blood leukocytes. Observed allele fractions based on whole-genome sequencing differed between twins, consistent with early post-zygotic segregation.

- a paternal allele with a mosaic pathogenic variant (c.1463del) or
- a paternal allele with a mosaic pathogenic variant (c.1466del) or
- a paternal allele with the wild-type sequence (no pathogenic variant).

Such a configuration is exceedingly unlikely, not due to the individual variants but because of their combination: two *de novo* variants arising in proximity on the same allele, each variant restricted to different cell lineages without occurring together in the same cell. We refer to this novel mutational configuration as “clustered monoallelic mosaicism” (cMoMa). Two explanatory models for the genesis of this genotype seem possible (Fig. 2A).

**Figure 2:**
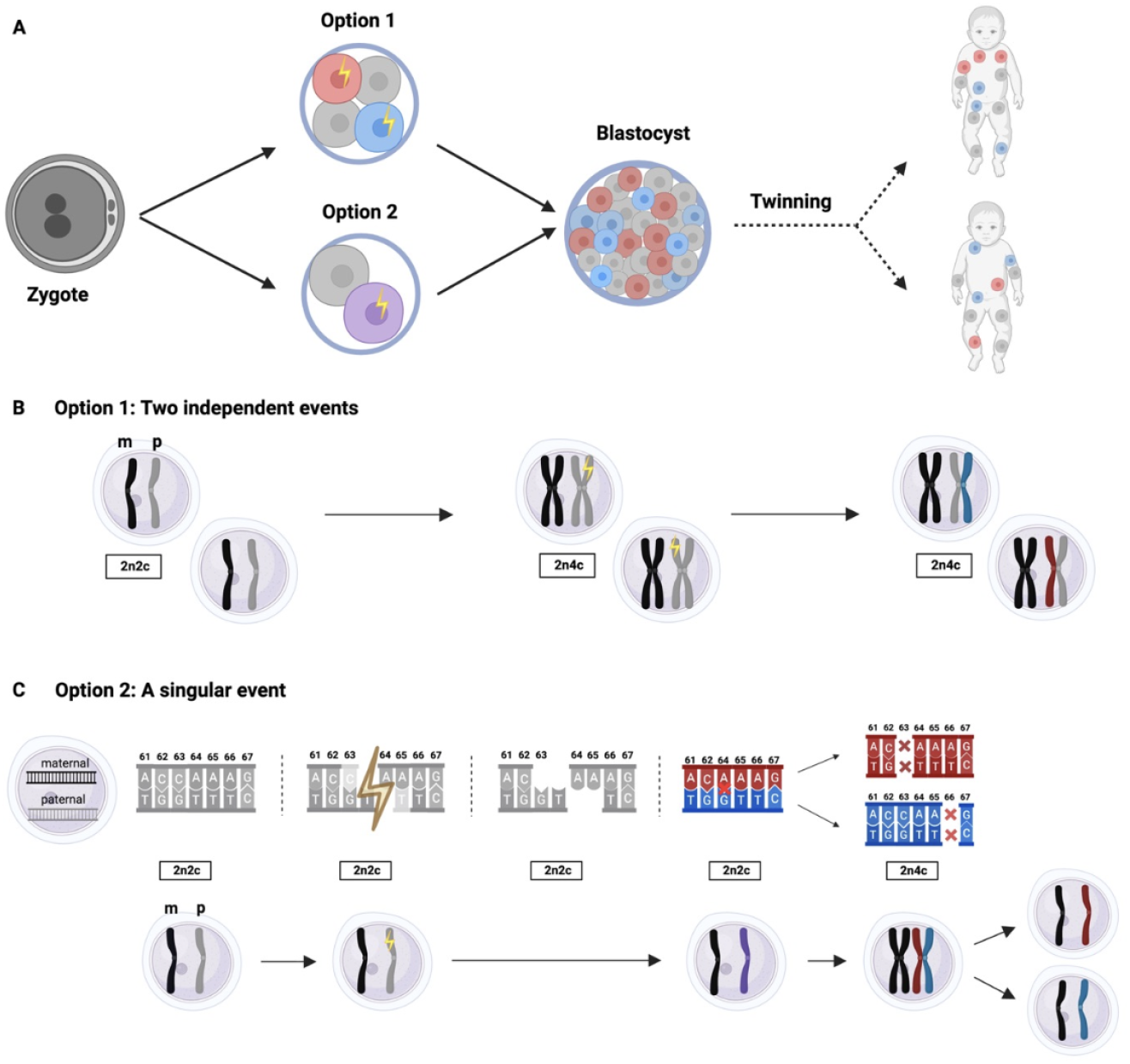
Mechanistic hypotheses for the origin of cMoMA. (A) Colored clones indicate lineage segregation through the blastocyst stage and into both twins. (B) Option 1: Two independent events. Two distinct *de novo* 1-bp deletions arise on the paternal allele in different early embryonic lineages. Because the events occur in separate cells, no double-mutant molecules are produced. Subsequent cleavages propagate two mutually exclusive mosaic lineages (red, blue). (C) Option 2: A singular event. A single mutational event (e.g. a double-strand break) occurs before sister-chromatid formation. Asymmetric processing during repair creates two distinct 1-bp deletions that are subsequently segregated to opposite sister chromatids after DNA replication.

**Model 1 (null hypothesis)** assumes that two independent mutational events produced the two 1-bp deletions. For both to reside on the same allele yet never co-occur on a single molecule, the deletions must have arisen in mutually exclusive embryonic lineages. Thus, the observed genotype can only be explained if the two deletions arose either in distinct early embryonic cells (Fig. 2B) or in the same cell, but on different sister chromatids, followed by segregation into separate lineages. Since both twins carry both variants, the events must have preceded twinning and occurred within the first approximately eight embryonic cell divisions.^4,5^ Under permissive assumptions for early embryonic mutation rates, the probability of two independent 1-bp deletions occurring within a few nucleotides during this period is vanishingly small (P <10^-7^, Supplemental S1).^6,7^ Given that the relatively high VAFs point to an even earlier embryonic origin, this estimate likely overstates the true likelihood of such a configuration arising independently.

Given the extremely low probability of the two-event scenario described in Model 1, this null hypothesis becomes statistically implausible. We thus considered whether a single mutational event could explain the observed genotype.

In **Model 2 (alternative hypothesis)**, a single mutational event arose during the earliest embryonic divisions, and subsequent error-prone repair gave rise to divergent sister-chromatid outcomes: a 1-bp deletion on one chromatid and a distinct, nearby 1-bp deletion on the other (Fig. 2C, Supplemental Fig. 4).

After cell division, this mechanism would be expected to yield daughter cells in a theoretical 1:1 ratio (Variant 1: Variant 2), thereby producing a mosaic pattern in which both variants are located on the same allele but never co-occur on the same sequencing read (i.e. are never found in the same cell, Fig. 1B). This mechanism explains the mutual exclusivity at the molecular level, the shared paternal origin by phasing, and the mosaic VAFs in leukocyte-derived DNA and DNA obtained from buccal swabs, with tissue-specific skew. Importantly, it does not rely on the occurrence of two highly improbable events in extremely close proximity.

Based on this mechanism, we would expect to observe similar cMoMas in large-scale datasets. An initial analysis of the COSMIC dataset (229,087 tumor samples) revealed an increased prevalence of clustered single-nucleotide deletions (Supplemental Fig. 3). However, long-read phasing data would have been required to accurately classify these events as true cMoMas. To address this, we additionally screened the MosaicBase dataset for individuals carrying distinct, same-gene variants located within 10 bp on the same allele.^8^ Two published cases met these criteria, each involving haplotype-resolved, mutually exclusive single-base deletions consistent with our cMoMa definition (Supplemental Table 1).^9,10^ All three cMoMa configurations, including the present case, consist of pairs of adjacent 1-bp deletions in short repetitive or structure-prone sequence motifs. In both *HNRNPU* and *WAS*, the deletions occur in short homopolymeric tracts, whereas in *ACVRL1*, they arise within a GC-rich region capable of forming secondary DNA structures. Moreover, in all three instances, the dual-deletion pattern was present across multiple tissues and likely occurred early during development. The recurrence of this mutational architecture suggests that cMoMa is not unique to our case and reflects a mechanism rather than an isolated anomaly.

## Discussion

Taken together, these findings position cMoMa as a mosaic counterpart to other clustered mutational phenomena: in cancer, *kataegis* describes localized substitution hypermutation; in the germline, multinucleotide mutations (MNMs) capture clusters of closely spaced mutations that occur more often than expected by chance.^7,11^ Engineered DSBs induced by Cas9 reproducibly yield heterogeneous, sequence-dependent indel spectra at cut sites, often including 1-bp deletions.^12^ Whereas other clustered mutational phenomena originate from *episodes* involving *multiple events*, cMoMa is more precisely defined by a *single event*. It is further characterized by lineage segregation: rather than co-occurrence within the same molecule, divergent outcomes are partitioned across sister chromatids and subsequently across descendant cell lineages, yielding mutually exclusive, monoallelic variants within one individual.

This configuration has immediate diagnostic implications. First, it illustrates that mosaicism at a locus does not necessarily reflect expansion of a single allele; instead, multiple distinct alleles can arise from a single event. Second, accurate recognition requires sequencing data with sufficient depth and phasing resolution to detect mutual exclusivity at the molecule level, determine parental origin, and distinguish true clustered variants from complex or recurrent artifacts. Third, for genetic counseling, recurrence risk equals the population baseline. Like other forms of mosaicism, cMoMas arise post-zygotically and are therefore incompatible with parental germline mosaicism or inherited transmission. Therefore, future children of the respective parents do *not* have an increased risk of recurrence. This point is particularly counterintuitive in twins: while shared disease in monozygotic twins is usually assumed to reflect inheritance, here both twins are affected by a post-zygotic event. Therefore, caution should be exercised to prevent misclassification as germline transmission, as it was the case in a previous cMoMa description.^9^

In summary, we define a previously unrecognized form of mosaicism likely caused by a single mutational event on one allele that gives rise to distinct pathogenic lineages through divergent sister-chromatid repair. We term this configuration “clustered monoallelic mosaicism” (cMoMa), defined by mutually exclusive, same-allele variants that never co-occur on a molecule. By indicating that a single mutational event may yield multiple lineage-segregated variants, cMoMa challenges the conventional assumption that one mutation produces one variant and extends the current list of categorizable genotypes, bridging mosaicism with other clustered mutation phenomena.

## Acknowledgements

The authors thank Laurine K. Sprehe, Samantha Sarli, and Tim Schubert for their intellectual insight and help in revising the manuscript. During the preparation of this work the authors used ChatGPT (OpenAI) in order to refine the language and improve clarity. After using this tool, the authors reviewed and edited the content as needed and take full responsibility for the content of the published article.

## Author contribution

JB* (Conceptualization, Data curation, Formal analysis, Investigation, Methodology, Project administration, Visualization, Writing - original draft, Writing - review & editing); JM* (Conceptualization, Data curation, Formal analysis, Investigation, Methodology, Project administration, Visualization, Writing - original draft, Writing - review & editing); JD* (Conceptualization, Data curation, Formal analysis, Investigation, Methodology, Project administration, Resources, Visualization, Writing - original draft, Writing - review & editing); SB (Resources, Writing - review & editing); LK (Formal analysis, Investigation, Supervision, Writing - review & editing); MB (Investigation, Writing - original draft); KG (Investigation, Writing - original draft); AKS (Formal analysis); THL (Formal analysis); KH (Resources, Supervision, Writing - review & editing); MH (Writing - review & editing); CS (Resources, Supervision, Writing - review & editing)

## Data Availability Statement

All data supporting the findings of this study are available within the article and its Supplementary Information files. Raw sequencing data (short-read WGS, long-read sequencing, and Sanger traces) are available from the corresponding author upon reasonable request, in accordance with institutional ethics approval and patient consent.

## Ethics Approval

This small study was approved by the ethics committee of the Medical Faculty Heidelberg (S-632/2023).

## Methods

### Whole-Genome Sequencing and Bioinformatic Analysis

Libraries were prepared from leukocyte-derived genomic DNA using the NEBNext® Ultra™ II FS DNA PCR-free Library Prep Kit for Illumina®, followed by high-throughput sequencing with 150-bp paired-end sequences on the NovaSeq 6000 system (Illumina). The data obtained from sequencing were bioinformatically analyzed using the varfeed™ pipeline, and the resulting variants were evaluated using the varvis® software from Limbus Medical Technologies GmbH (v. 2.1.0) with a mean coverage of 46. All annotations refer to the human reference genome GRCh38 (hg38). Various prediction programs, some of which are integrated into varvis®, were consulted to assess the variants. These include, for example, Fathmm, MetaLr, MetaSvm, MutAssessor, MutTaster, ScSnvAda, ScSnvRf, and SIFT. Information from public databases such as ClinVar, HGMD® Professional, OMIM, UCSC, Decipher, and the Database of Genomic Variants (DGV) was also used. The nomenclature of the Human Genome Variation Society (HGVS; http://www.hgvs.org/mutnomen/) is used to classify variants. Pathogenicity is classified based on the ACMG and IARC guidelines.^13,14^

### Sanger Sequencing

Genomic DNA extracted from buccal mucosa and from leukocytes of the twins was used for PCR amplification and further sequencing of exon 7 of the *HNRNPU* gene (NM_031844.3) comprising both deletions in question (primers on request). This method has a sensitivity of 97-99% for identifying sequence alterations in the region of interest.

### Long-Read Sequencing

Long-read sequencing was performed using the Oxford Nanopore platform. Library preparation was conducted using the Ligation Sequencing Kit V14 (Oxford Nanopore Technologies, SQK-LSK114-XL) according to the manufacturer’s instructions, with modifications based on the Rapid CNS2 approach. Briefly, 2·5-3 µg of DNA were sheared to an average fragment length of 25 kb using Covaris g-TUBEs (Covaris, 520079). During the end-prep step, the AMPure XP beads ratio was adjusted to 70 µl. Adapter ligation was performed with 10 minutes of mixing on a Hula Mixer, and during the final elution step samples were incubated for 3 minutes at 37 °C. Subsequently, 600-700 ng of the prepared library was loaded onto a new MinION Flow Cell R10.4.1 (ONT, FLO-MIN114) and sequenced for 24 hours with enabled adaptive sampling that limited data acquisition to sequences of the *HNRNPU* locus ± 20 kb. Bioinformatic analysis was carried out using the Rapid-CNS2 pipeline.

### Cosmic Analysis

Variants from the COSMIC tumor dataset (229,087 samples; 1,048,574 variants) were filtered to include single-nucleotide deletions only. Variant clusters were defined per sample and gene, and only clusters with inter-variant distances <100 bp were retained (n = 59). Prevalence was plotted as a histogram using a sliding window (max_distance = 100 bp; window = 3 bp; step = 2 bp). A kernel density estimate (bandwidth = 2) was overlaid, and a Poisson-based expected distribution (λ = 0·8) was added for comparison. All analyses and visualizations were performed in Python.

### MosaicBase Analysis

We reanalyzed MosaicBase to identify candidate clustered monoallelic mosaicism (cMoMa) events.^8^ Entries corresponding to single-cell sequencing datasets (disease: Cockayne syndrome, Xeroderma pigmentosum, asymptomatic, human skin fibroblasts, or NA) were excluded, as these studies lacked haplotype resolution required for cMoMa identification. For the remaining entries, we searched for individuals carrying ≥2 distinct variants in the same gene located within ≤10 bp. Redundant or identical calls were collapsed, and distinct variant pairs were reported as cMoMa candidates. Two published haplotype-resolved single-base deletions met these criteria.^9,10^

