## Supplements for "Too rare to be random: genetic finding suggests previously unrecognized path of mutagenesis"

### cMoMa Calculations

#### 1 A null model for near pairs under Poisson event counts

We model the number $N$ of early 1-bp deletions as Poisson with mean $\lambda$. Conditional on $N = n$, the $n$ deletion coordinates are assumed to be independent and uniformly random over the diploid genome $\{1, \dots, G\}$.[1]

We call two events a *near pair* if their coordinates differ by at most $w/2$ bases, i.e. they lie within a symmetric window of total length $w$ around an index site. Here we set $w = 6$ bp to represent the window $\pm 3$ bp *excluding* the index base (distinct positions).

**General result (used throughout).** For $N \sim \text{Poisson}(\lambda)$ and $w \ll G$, the expected number of near pairs is derived as follows.

Given $N = n$ deletions, there are $n(n-1)/2$ possible pairs, and each pair has probability about $w/G$ of being within the same local window. Therefore,

$$\mathbb{E}[\#\text{near pairs} \,|\, N = n] = \frac{n(n-1)}{2} \cdot \frac{w}{G}.$$

Taking the expectation over $N$ gives

$$\mathbb{E}[\#\text{near pairs}] = \mathbb{E}\Big[\frac{N(N-1)}{2} \cdot \frac{w}{G}\Big] = \frac{w}{2G}\,\mathbb{E}[N(N-1)].$$

For any random variable, $\mathbb{E}[N(N-1)] = \mathbb{E}[N^2] - \mathbb{E}[N]$. Using $\text{Var}(N) = \mathbb{E}[N^2] - (\mathbb{E}[N])^2$, we can write

$$\mathbb{E}[N(N-1)] = \text{Var}(N) + (\mathbb{E}[N])^2 - \mathbb{E}[N].$$

For a Poisson variable, $\text{Var}(N) = \mathbb{E}[N] = \lambda$, so

$$\mathbb{E}[N(N-1)] = \lambda + \lambda^2 - \lambda = \lambda^2.$$

Hence,

$$\boxed{\mu_{\text{near}} = \frac{\lambda^2}{2} \cdot \frac{w}{G}.}$$

This is the expected number of near-pair events under the null model of independent, uniformly distributed deletions.

#### 2 Early scenario restricted to divisions 2 and 3 (2→4 and 4→8 cells)

**Rationale.** A first-cleavage (zygote) event would eliminate the wild-type paternal lineage, which is inconsistent with the observed VAFs; therefore we restrict the early scenario to the *second and third* cleavages.

**Assumptions.** Diploid genome size $G \approx 6.4 \times 10^9$ bp and window $w = 6$ bp. For the first three cleavages, Chapman et al. estimate $\approx 2.4$ SNVs per daughter cell per division [1]. We convert SNVs to 1-bp deletions (1-bp dels) using an indel:SNV ratio of 1:13.78 and a 1-bp deletion fraction among indels of 33.8% [2].

##### Expected number of 1-bp deletions and near pairs

Divisions 2 and 3 produce $4 + 8 = 12$ daughter cells, giving

$$S_{(2,3)} = 12 \times 2.4 = 28.8 \text{ SNVs}.$$

[1] This simplified null model assumes that each deletion arises independently and with equal probability across the diploid genome, ignoring local sequence-context biases. It thus provides a baseline estimate of how rare such events would be under independence.

Converting to 1-bp deletions yields the Poisson mean

$$\lambda_{(2,3)} = 28.8 \times \frac{1}{13.78} \times 0.338 = 28.8 \times 0.07257 \times 0.338 \approx \mathbf{0.7064}.$$

Thus

$$\mu_{\text{near}} = \frac{\lambda_{(2,3)}^2}{2} \cdot \frac{w}{G} = \frac{0.7064^2}{2} \cdot \frac{6}{6.4 \times 10^9} \approx \mathbf{2.34 \times 10^{-10}}.$$

#### 3 Aggregating through the 8th mitosis (twinning window)

**Assumptions.** We aggregate across the first eight mitotic divisions (to $\sim$ 256 cells). For divisions 1–3 we use 2.4 SNVs per daughter per division; for divisions 4–8, we use $<$ 0.9 SNVs per daughter per division. The same indel:SNV and 1-bp deletion fractions apply.

##### Expected number of 1-bp deletions and near pairs

**Divisions 1–3:** $2 + 4 + 8 = 14$ daughters $\Rightarrow$ $14 \times 2.4 = 33.6$ SNVs.
**Divisions 4–8:** $16 + 32 + 64 + 128 + 256 = 496$ daughters $\Rightarrow$ $496 \times 0.9 \approx 446.4$ SNVs.
**Total:** $S \approx 33.6 + 446.4 = 480$ SNVs.

Converting to 1-bp deletions:

$$\lambda_{(\leq 8)} = 480 \times \frac{1}{13.78} \times 0.338 \approx 480 \times 0.07257 \times 0.338 \approx \mathbf{11.8}.$$

Hence,

$$\mu_{\text{near}} = \frac{\lambda_{(\leq 8)}^2}{2} \cdot \frac{w}{G} = \frac{11.8^2}{2} \cdot \frac{6}{6.4 \times 10^9} \approx \mathbf{6.53 \times 10^{-8}}.$$

**Interpretation.** In a simplified model that assumes deletions occur independently, are uniformly distributed across the diploid genome, and follow Poisson-distributed event counts, the expected number of near-pair deletions within $\pm 3$ bp is exceedingly small. When considering only the second and third embryonic cleavages, this expectation is on the order of $10^{-10}$. Even when extending the calculation to include all events through the eighth mitosis, the expected near-pair count rises only to about $10^{-8}$. These estimates indicate that two closely spaced 1-bp deletions are highly unlikely to result from independent mutational events under this model.

#### Supplementary Material

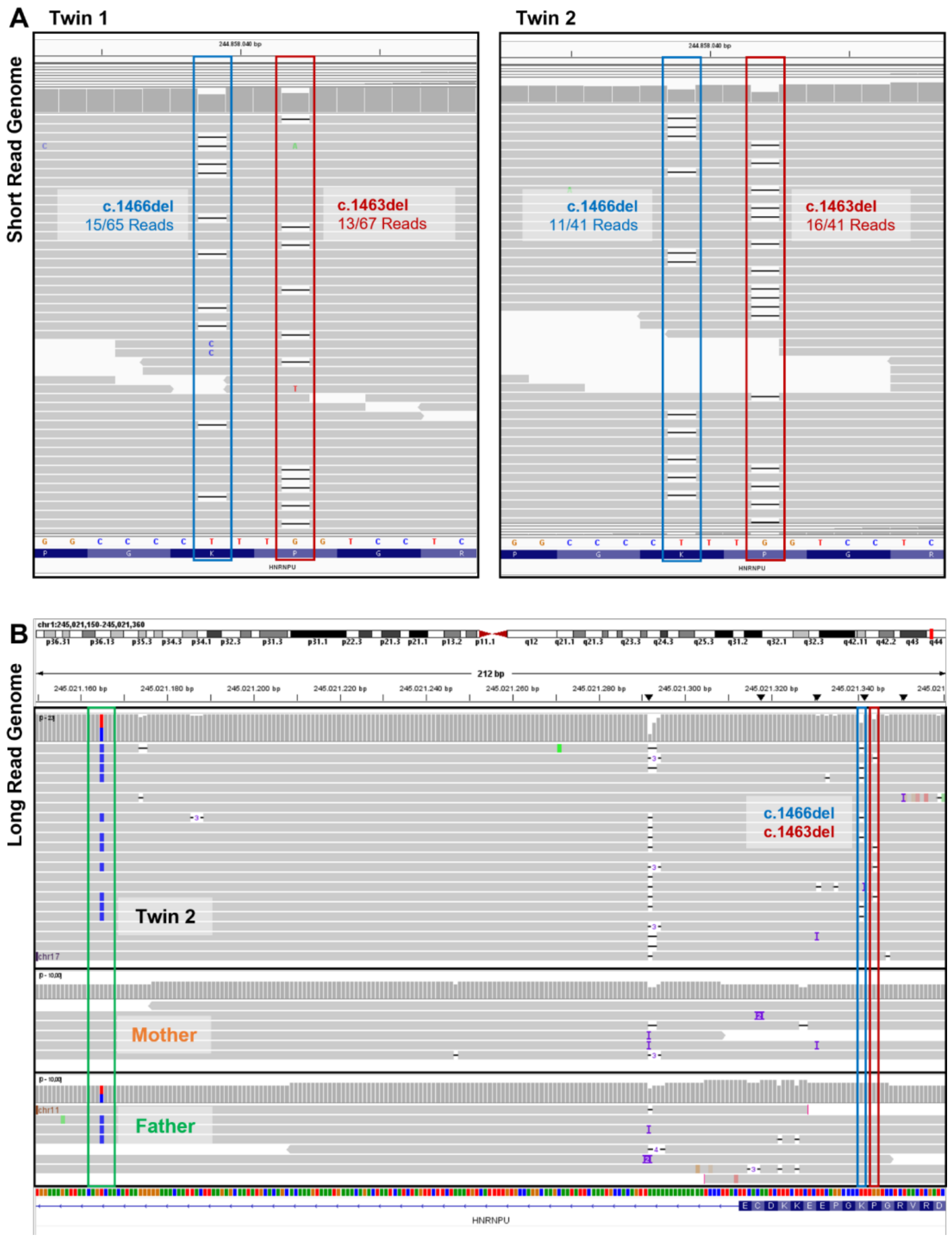


**Supplemental Figure 1: Whole-genome sequencing of the leukocyte-derived DNA**

Short-read whole-genome sequencing of leukocyte-derived DNA from the twins and their parents identified two closely spaced *de novo* 1-bp deletions in *HNRNPU* (NM_031844.3; c.1463del and c.1466del) on chromosome 1 (A). They were present in mosaic state with variant allele frequencies (VAFs) ranging from ~20-39%. Short-read genome sequencing data demonstrated that both deletions were never observed on the same read (A), nanopore long-read sequencing was used to determine the allelic phase, revealing both on the paternal allele, indicated by a paternal SNP (green box) on the same read as both deletions (blue/red box) (B).

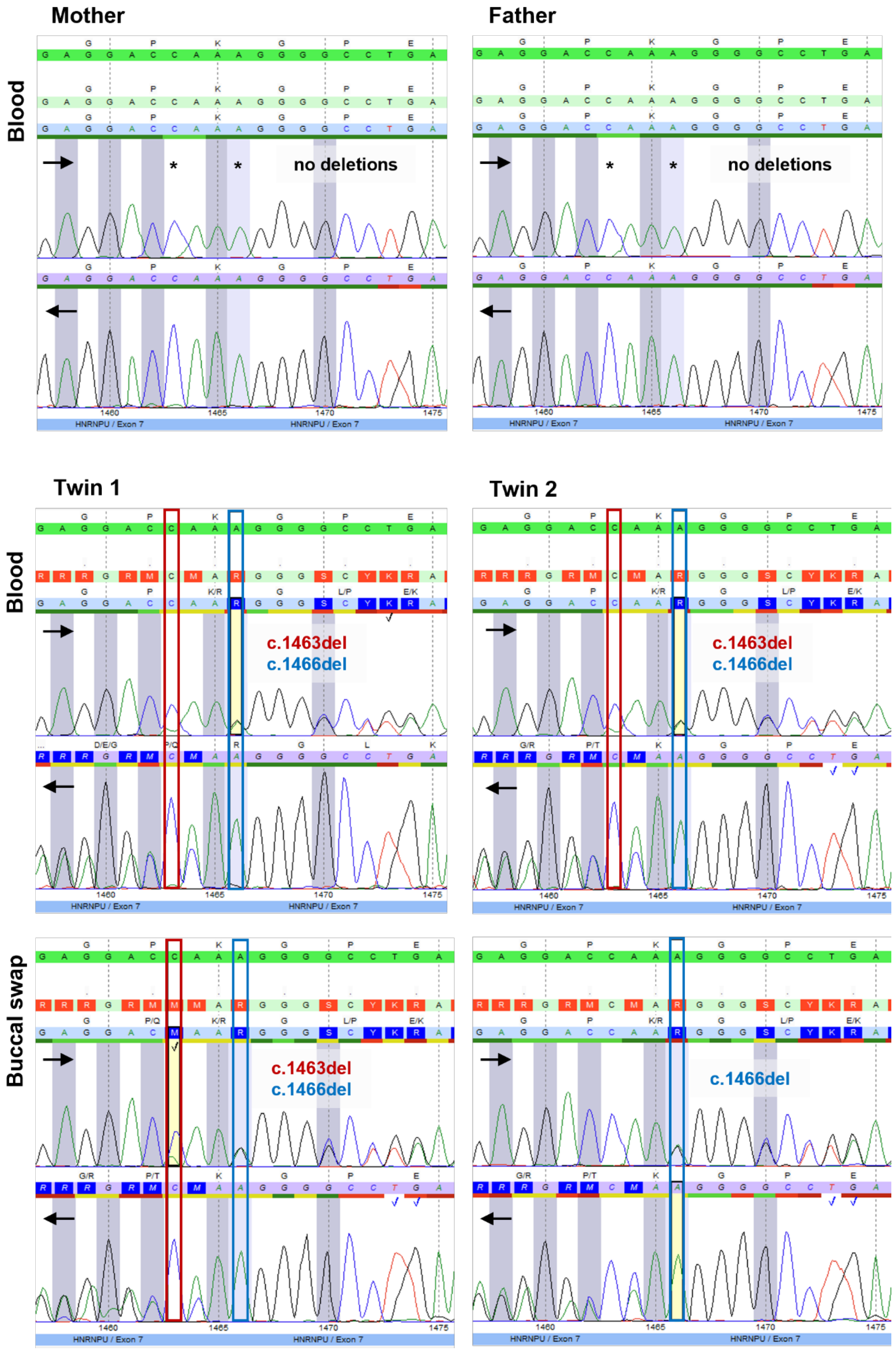


**Supplemental Figure 2: Sanger sequencing of leukocyte-derived and buccal-derived DNA**

In twin 1, both variants (*HNRNPU* NM_031844.3; c.1463del and c.1466del) were present in mosaic form in both tissues. In contrast, twin 2 exhibited both deletions in mosaic form only in leukocyte-derived DNA, while only one heterozygous deletion (c.1466del) was detected in the buccal swab, consistent with lineage skew and assay sensitivity limits.

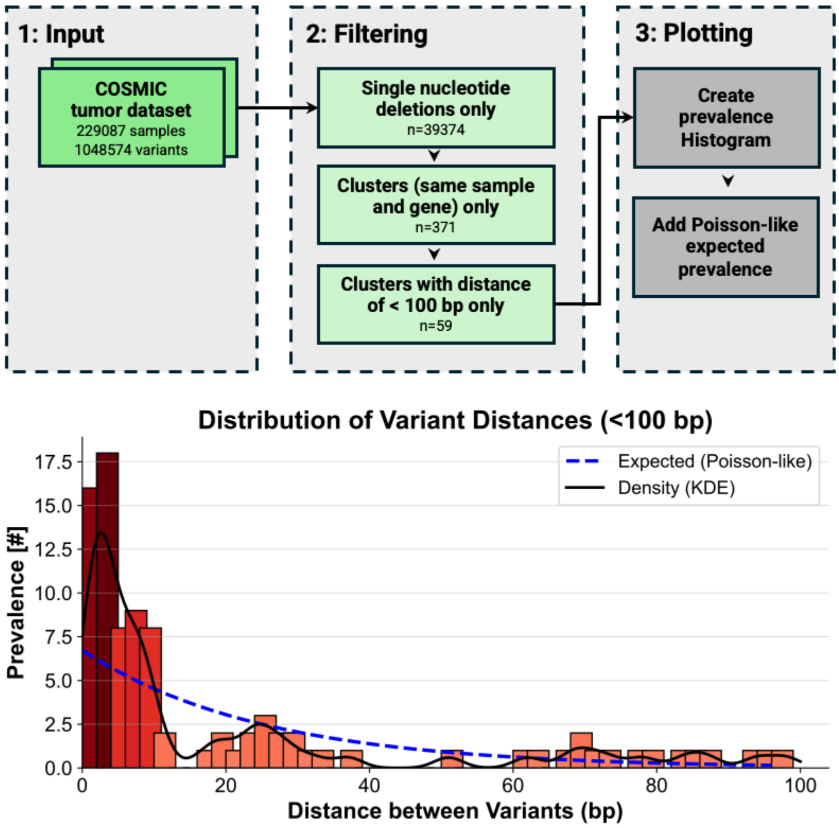


**Supplemental Figure 3: Prevalence of clustered single nucleotide deletions in COSMIC dataset**

This figure shows the *in silico* analysis pipeline (input, filtering, and plotting) and the resulting prevalence of clustered single-nucleotide deletions in the COSMIC dataset. For each sample-variant pair, where two variants were located within 100 bp of each other, we calculated the prevalence (y-axis) as a function of inter-variant distance (x-axis). Bars represent sliding-window binning (max_distance = 100, window_size = 3, step_size = 2) and are additionally color-coded by height. The corresponding density curve (black line) was derived using a kernel density estimate (KDE) with a bandwidth factor of 2. The dashed blue line indicates the expected distribution based on a Poisson model ($\lambda$ = 0.8, equal to the mean of the distances). The figure highlights the deviation between the observed and mathematically expected distributions.

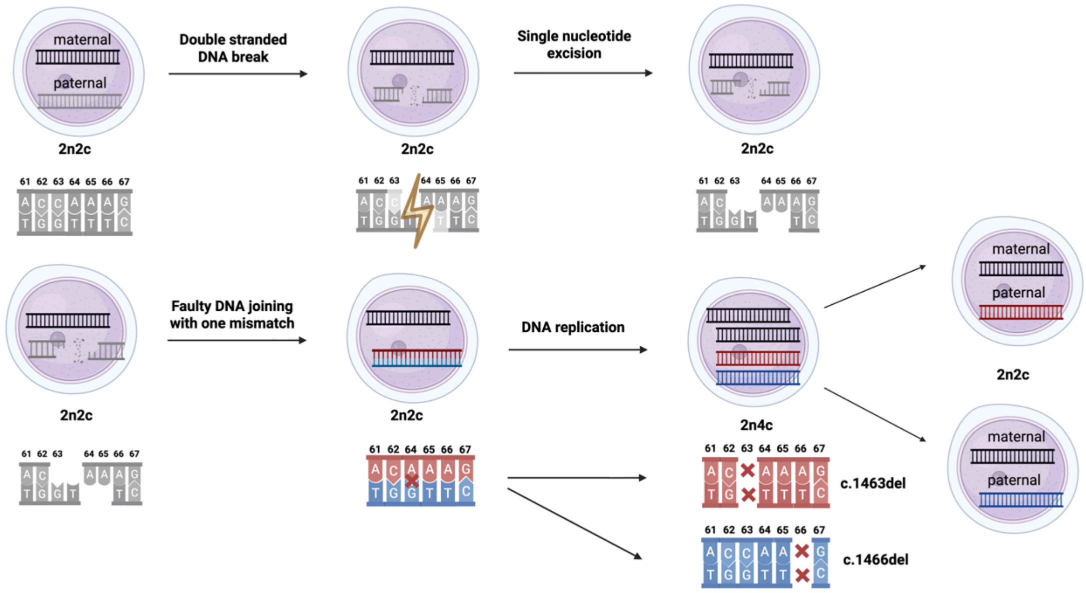


**Supplemental Figure 4: Proposed molecular mechanism underlying cMoMa formation**

Schematic representation of a potential mechanism of cMoMa formation: A DSB arises on the paternal allele, followed by end processing and single-nucleotide loss on each break end. Re-ligation of the processed fragments introduces a transient base-pair mismatch, producing two subtly different sister chromatids. After replication, the divergent chromatids segregate into separate daughter cells, yielding two independent mosaic lineages that together recapitulate the genotype observed in this study.

**Supplemental Table 1: Overview of the cMoMa cases presented in this study**

| Study | This study | Eyries et al., 2012 | Dobbs et al., 2007 |
|---|---|---|---|
| **Gene Name** | *HNRNPU* | *ACVRL1* | *WAS* |
| **Variant 1 (HGVS)** | c.1463del | c.1388del (p.Gly463Alafs*2); within G(2) tract | c.755del (within C(3) tract c.755–c.757; codon 241) |
| **Variant 2 (HGVS)** | c.1464del (within A(3) tract c.1464–c.1466) | c.1390del (p.Leu464*); within C(2) tract) | c.758del (within C/A homopolymer; codon 242) |
| **Distance (bp)** | 1–3 bp (adjacent, within homopolymer) | 1-4 bp (adjacent, within homopolymers) | 1–3 bp (adjacent, within homopolymer) |
| **Sequence Context** | Located within an A/T-rich repetitive region | Local hairpin structure | Short run of cytosines and a brief palindromic motif |
| **Variant type** | 1-bp deletions | 1-bp deletions | 1-bp deletions |
| **Affected individual** | Monozygotic twins | Single female with HHT + PAH | Single carrier female (II:2) for WAS |
| **Evidence for same haplotype / mutual exclusion** | Long-read phasing shows mutually exclusive deletions on the same paternal allele | Microsatellite haplotyping; same maternal haplotype. | Allele-specific PCR: both deletions on same great-grandpaternal haplotype |
| **Notes** | Detected in blood and buccal DNA; no double-mutant reads; early embryonic origin. | Early embryonic origin. | Authors correctly identified the dual-deletion genotype but misinterpreted the origin as "bichromatid mutation in a male gamete"; the mosaic distribution indicates a cMoMa event. Germline mosaicism, since each variant was passed on. |
